# Systematic review search strategies are poorly described and not reproducible: a cross-sectional meta-research study

**DOI:** 10.1101/2023.05.11.23289873

**Authors:** Melissa L. Rethlefsen, Tara J. Brigham, Carrie Price, David Moher, Lex M. Bouter, Jamie J. Kirkham, Sara Schroter, Maurice P. Zeegers

## Abstract

**Objective:** To determine the reproducibility of biomedical systematic review search strategies.

**Design:** Cross-sectional meta-research study.

**Population:** Random sample of 100 systematic reviews indexed in MEDLINE in November 2021.

**Main Outcome Measures:** The primary outcome measure is the percentage of systematic reviews for which all database searches can be reproduced. This was operationalized as fulfilling six key PRISMA-S reporting guideline items (database name, multi-database searching, full search strategies, limits and restrictions, date(s) of searches, and total records) and having all database searches reproduced within 10% of the number of original results.

**Results:** The 100 systematic review articles contained 453 database searches. Of those, 214 (47.2%) provided complete database information (named the database and platform; PRISMA-S item 1). Only 22 (4.9%) database searches reported all six PRISMA-S items. Forty-seven (10.4%) database searches could be reproduced within 10% of the number of results from the original search; 6 searches differed by more than 1000% between the originally reported number of results and the reproduction. Only one systematic review article provided the necessary details for all database searches to be fully reproducible.

**Conclusion:** Systematic review search reporting is poor. As systematic reviews and clinical practice guidelines based upon them continue to proliferate, so does research waste. To correct this will require a multi-faceted response from systematic review authors, peer reviewers, journal editors, and database providers.

## INTRODUCTION

Systematic reviews provide synthesized evidence using robust, pre-specified methods to reduce bias and enable better decision-making by health care professionals, patients, and policy makers.[1, 2] Transparent reporting is essential because it enables readers to evaluate the value of systematic reviews and to identify potential sources of bias that impact the review’s findings.[3] A sensitive literature search encompassing multiple information sources is central to the methodology of systematic reviews.[1, 2] The search must be transparently and completely reported. It should be possible to reproduce the search and to assess whether this leads to similar results.

Systematic reviews are often poorly reported, which impacts reproducibility and leaves readers unable to assess potential biases.[4-7] Systematic review searches are particularly vulnerable to incomplete reporting.[8-12] Estimates of systematic review search reproducibility vary widely, in part because many researchers have developed their own criteria for estimating reproducibility.[9, 10, 13-15] In 2021, PRISMA-S, an extension of PRISMA, a guideline for reporting systematic reviews, was designed to help systematic review teams report their searches for maximum transparency and reproducibility.[16] PRISMA-S offers a standard for the details systematic review searches need to describe to be reproducible.[16] However, this is not enough to determine actual search reproducibility; reproducing the search is required.

The objective of this study was to understand the reproducibility of systematic review searches by examining the completeness of reporting and reproducing each search. Understanding the reproducibility of published searches can serve as a baseline for improvement.

## METHODS

Prior to data collection, the study protocol was registered on the Open Science Framework (OSF).[19] The registration was amended during the study to reflect minor changes in the protocol (see study protocol amendments section).

### Identification and selection of articles

We aimed for a cohort of 100 systematic reviews. We first searched for systematic reviews indexed in MEDLINE in one calendar month, November 2021. Because it can take 6 months or more to be indexed in MEDLINE, this enabled us to get a snapshot of items from a range of publication dates. We only included publications in English for pragmatic reasons. We adapted the search published in Page et al[7] and conducted it on December 20, 2021, using Ovid MEDLINE ALL <1946 to December 17, 2021>; the complete search strategy (https://osf.io/4mcgu) is available in the study’s OSF Project site.[20] Results were deduplicated using Covidence.[21]

To be considered a systematic review, articles needed to meet the PRISMA-P definition of a systematic review,[22] namely they must explicitly state methods to locate studies (i.e., the search), methods to select studies (i.e., screening using eligibility criteria), and methods to synthesize the studies, whether qualitatively or quantitatively. In addition, the article had to explicitly state that one or more literature databases (e.g., MEDLINE) were searched as part of the methods. These database searches must have occurred in 2020 or later, as indicated in the article text or supplementary materials. We excluded articles that solely searched PubMed prior to May 2020,[23] due to the structural changes to the PubMed database in May 2020.[24] We excluded scoping reviews, evidence maps, and other review types that use iterative search methods. We had no restrictions to the study design and the study question of the primary studies the systematic review had included.

Titles and abstracts were screened in duplicate using Covidence[21] to assess whether articles appeared to meet the definition of a systematic review. If at least one reviewer agreed that the studies met the eligibility criteria, they were included. This liberal screening method, as used in Page et al.,[7] helps to account for the difficulty in determining whether something is a “true” systematic review, as no single definition of a systematic review exists. After the title/abstract screening, we randomly ordered the remaining articles using Microsoft Excel’s RAND function syntax. We reviewed the full text of studies in the randomly ordered list in duplicate using all inclusion and exclusion criteria until 100 studies were identified for inclusion. Conflicts were resolved through discussion between the two reviewers and a third reviewer where needed. We opted to use a sample of 100 articles to provide a workable set for database search reproductions.

### Data collection: data extraction phase

Data from each article in the sample were extracted in duplicate using a custom extraction form in Covidence.[21] Prior to beginning data extraction, reviewers trained together using a set of systematic reviews outside the study sample to ensure there was a shared understanding of each data extraction field. A copy of the complete data extraction form (https://osf.io/gm74t) is available in the project’s OSF repository.[20] Pairs of reviewers extracted information on the details of the search and search reporting (e.g., databases searched, number of results per database, database search dates, use of limits/restrictions, presence of a PRISMA flow diagram, etc.) and adherence to the six PRISMA-S items noted in Table S1 (https://osf.io/g6prk). The six PRISMA-S criteria included those directly related to database searches: database name (Item 1), multi-database searching (Item 2), full search strategies (Item 8), limits and restrictions (Item 9), dates of searches (Item 13), and total records (Item 15). Data was extracted from the published article and supplementary materials; protocols were not considered. Consensus was achieved through discussion where necessary. Using the extracted data, we calculated database search and systematic review search adherence to the 6 PRISMA-S items.

### Data collection: reproduction stage

To conduct the search reproductions and to extract data relating to the reproductions, one assessor (MLR) extracted data and conducted the initial reproduction. A second assessor (TJB, CP, or JR) validated the data and reproduction. To conduct the reproduction, we used the same databases and platforms as used by the original systematic review teams used, where known. At this stage, we excluded database searches conducted in Japanese and Chinese platforms that required non-Latin characters or operated differently depending on geographical location. We also excluded database searches when we were unable to locate an assessor with access to the database and platform specified. Each search was reproduced by copying and pasting the search directly from the original article when possible. We also reproduced searches that were incomplete, but were described well enough to reconstruct a full search strategy. We anticipated that expertise would be required to reproduce many searches. If it was necessary to apply expert knowledge or if errors were evident and required fixing, these interventions were undertaken and noted.

We applied the same database limits as the original search, and searches were restricted to database records entered on or prior to the date of the last search, as reported by the systematic review authors. If only the month was reported, the search was re-executed with a date limit set on the last day of the reported month. Estimated search dates were used for any search date that was unclearly reported. For databases or platforms without the capacity to limit by database entry date, publication date limits were applied to remove recent records from the search results.

The assessor captured the details of the original search, plus details of the reproduced search, including assumptions, the complete reproduction search strategy, notes on where expert knowledge was applied, details of the database and platform used, search date, and number of results retrieved. In addition, we captured screenshots of each search for additional records. Second assessors added additional observations. Standardized data was captured using a Qualtrics form (https://osf.io/f97hc) after the completion of each reproduction and validation.[20] If there was not enough detail to reproduce a search, even with expert knowledge, the search was considered impossible to reproduce.

### Primary and secondary outcomes

For the purposes of this study, we considered a **database search** to be reproducible if it meets six PRISMA-S items, can be re-executed without editing in the named database and platform,[17] and the number of results retrieved is within 10% of the original search results. We considered a **systematic review search** to be reproducible if these elements are met for *all databases* searched. The primary outcome of this study is the percentage of systematic reviews for which all database searches can be fully reproduced. Secondary outcomes include:

1. The percentage of systematic reviews for which
  a. one database search can be fully reproduced
  b. more than one database search can be fully reproduced
2. The percentage difference between the number of search results reported in a systematic review versus the number of results in the reproduced search
3. The number of systematic reviews meeting individual elements needed for reproducibility for one or more databases

For the purposes of our study, we used the definition of “reproducibility” from the National Academies of Science, Engineering, and Medicine, “Reproducibility is obtaining consistent results using the same input data; computational steps, methods, and code; and conditions of analysis.”[18]

### Data analyses

To test for differences between the original search (as reported) and the reproduced search, we determined how many records we retrieved versus how many had been previously reported. We calculated the percentage difference between the original and reproduction searches (difference in number of results identified / number of results identified by the original search X 100%). If expert knowledge was required to re-run the search, we did not calculate the difference due to our inability to assess whether the reproduction was accurate. For many database searches, the number of results per database was not reported; for these, we could not assess the difference between the initial search and its reproduction. We used a 10% or less difference in results as an indicator of reproducibility to allow for known variability of database results over time and the potential for unclear search date reporting.[25-27]

### Patient and public involvement

No patients or members of the public were directly involved in the design, conduct, or analysis of this study. Our primary audience is researchers, peer reviewers, and editors, but knowing that biased systematic reviews, especially those that serve as the basis of clinical practice guidelines, can impact patient care inspired this work.

### Study protocol amendments

Minor changes were made to data collection elements between protocol registration and data collection. We clarified when database searches would not be reproduced. In addition, we transitioned to providing search reproduction documentation notes on some variables instead of capturing standardized data. During the analysis, we added an additional secondary outcome, the number of database searches meeting individual elements needed for reproducibility, as collecting this data was required to fulfill the other outcomes. The final protocol is available on the OSF Project site.[20] We conducted a post-hoc sensitivity analysis to analyze whether changing the required percentage difference (pre-determined at 10%) would alter results of our primary outcome (number of reproducible systematic reviews) or secondary outcome (number of database searches meeting individual elements needed for reproducibility).

## RESULTS

The search retrieved 8,905 results; after removing duplicates, 8,640 results remained for title/abstract screening. 4,124 systematic review articles remained after title/abstract screening and were randomly ordered for full-text screening. One hundred and sixty-three articles were reviewed in full text before 100 articles that met all eligibility criteria were identified. The 100 articles represented 78 different journals (Table S2 (https://osf.io/skrb3)[20]). All articles except one were published in 2021. Most reported searching three or more databases (91%; 91/100) and presented PRISMA flow diagrams (99%; 99/100). 23% (23/100) did not provide a search strategy. 44% (44/100) had a registered or published study protocol (Table S3 (https://osf.io/vabpj)[20]).

### Reporting of PRISMA-S items

The final set of 100 systematic reviews contained 453 database searches (range: 1-14 databases; median: 4 databases per article). Of those, complete database information, including naming the database and platform (PRISMA-S item 1), was available for 47.2% (214/453) (Table 1). Only 4.9% (22/453) database searches clearly reported all six PRISMA-S items. Least commonly reported were item 9, limits and restrictions, and item 13, dates of searches. Limits and restrictions were fully reported for 22.1% (100/453) of database searches, and the exact date of the search was provided for 22.7% (103/453) database searches.

**Table 1.** Number and percentage of database searches and systematic review searches meeting criteria, as well as the number of systematic review searches with one or more database searches meeting criteria. There were 453 total database searches in 100 systematic review searches. For example, 22 systematic reviews had one or more database searches with reproduction results within 10% of the original, but only one systematic review search had all database searches with reproduction results within 10% of the original.

|  | Database searches meeting criteria (n=453) |  | Systematic reviews with one or more database searches meeting criteria (n=100) |  | Systematic review searches meeting criteria (n=100) |  |
| --- | --- | --- | --- | --- | --- | --- |
|  | n | % | n | % | n | % |
| **PRISMA-S Items** |  |  |  |  |  |  |
| Item 1: Database name | 214 | 47.2% | 90 | 90.0% | 9 | 9.0% |
| Item 2: Multi-database searching | 368 | 81.2% | 99 | 99.0% | 45 | 45.0% |
| Item 8: Full search strategies | 133 | 29.4% | 40 | 40.0% | 15 | 15.0% |
| Item 9: Limits and restrictions | 100 | 22.1% | 21 | 21.0% | 20 | 20.0% |
| Item 13: Dates of searches | 103 | 22.7% | 25 | 25.0% | 19 | 19.0% |
| Item 15: Total records | 226 | 49.9% | 58 | 58.0% | 43 | 43.0% |
| **All 6 PRISMA-S Items** | **22** | **4.9%** | **6** | **6.0%** | **2** | **2.0%** |
| **Reproducibility** |  |  |  |  |  |  |
| Reproduction results within 10% of original | 47 | 10.4% | 22 | 22.0% | 1 | 1.0% |
| **Total** |  |  |  |  |  |  |
| **All 7 criteria (PRISMA-S and Reproduction within 10% of original)** | **16** | **3.5%** | **6** | **6.0%** | **1** | **1.0%** |

Six (6%; 6/100) systematic reviews clearly reported all six PRISMA-S items for at least one database search (Table 1) and five of these six systematic reviews reported all PRISMA-S items for more than one database. Only two (2%; 2/100) systematic reviews fully reported all six PRISMA-S items for all of their searched databases. The most commonly fully reported PRISMA-S item was item 2 (multi-database searching) for which either a “yes” or “not applicable” response was considered meeting criteria (Table S1 (https://osf.io/g6prk)[20]). “Not applicable,” which meant that they did not conduct a search in a platform where multiple databases could be searched simultaneously, applied to 41 of the 45 systematic reviews, leaving only four systematic reviews which clearly reported conducting a multi-database search. The second most commonly reported was item 15, total records, where 43 (43%; 43/100) articles reported the number of results for each database searched for the systematic review.

Conversely, only nine (9%; 9/100) systematic reviews reported item 1 (database name) for all databases searched.

### Reproduction

Of the 453 database searches, 64.2% (291/453) provided enough information for us to attempt a reproduction. We were unable to run 3.8% (17/453) of the searches we attempted to reproduce, leaving 60.5% (274/453) remaining searches to execute (see Figure 1). 31.3% (142/453) searches required expert knowledge to recreate, most commonly selecting the platform to use (70.4%; 100/142). 64.1% (91/142) of the searches requiring expert knowledge needed intervention in multiple aspects of the search. These interventions included selecting a platform or database(s); fixing, adapting, or adding Boolean logic; adding limitations; fixing search syntax; selecting fields to search; and/or other changes. Detailed notes, including descriptions of all interventions taken, and screenshots for each attempted and/or completed reproduction are available on the OSF Project site.[20]

**Figure 1.**
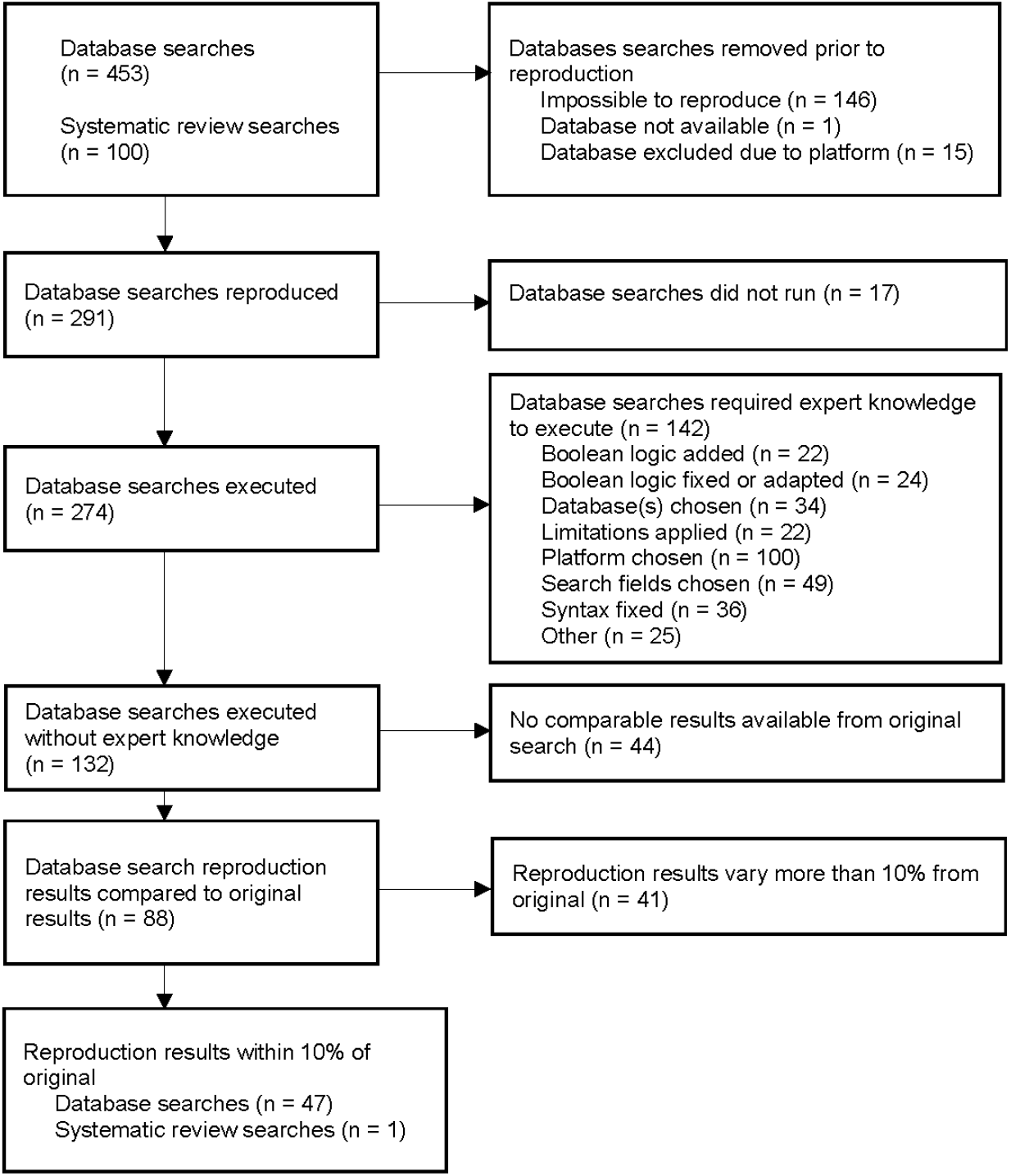
Flow Diagram of Database Search Reproductions

Results were not reported for all database searches. We were able to compare the number of results of 88 reproduced searches (from 39 systematic reviews) to the number of results retrieved by the original database searches. Forty-seven (10.4%; 47/453) database searches were able to be reproduced within 10% of the number of results from the original search; 3 (0.7%; 3/453) of those reproductions matched the results numbers exactly. For 41 database search reproductions, the results varied more than 10% between original and reproduction. Six of these searches differed by more than 1000% between the originally reported number of results and the reproduction, though most (73.2%; 30/41) varied positively or negatively between 10.1%-100.0% (see Figure 2). 22.0% (22/100) of systematic reviews had one or more database searches that were able to be reproduced within 10% of the originally reported results, and only one systematic review[28] had all of its database searches reproduced within 10% of the originally reported results (Table 1). This paper by Nguyen et al. also fully reported all 6 PRISMA-S items, thus being the only fully reproducible systematic review in our study.[28]

**Figure 2.**
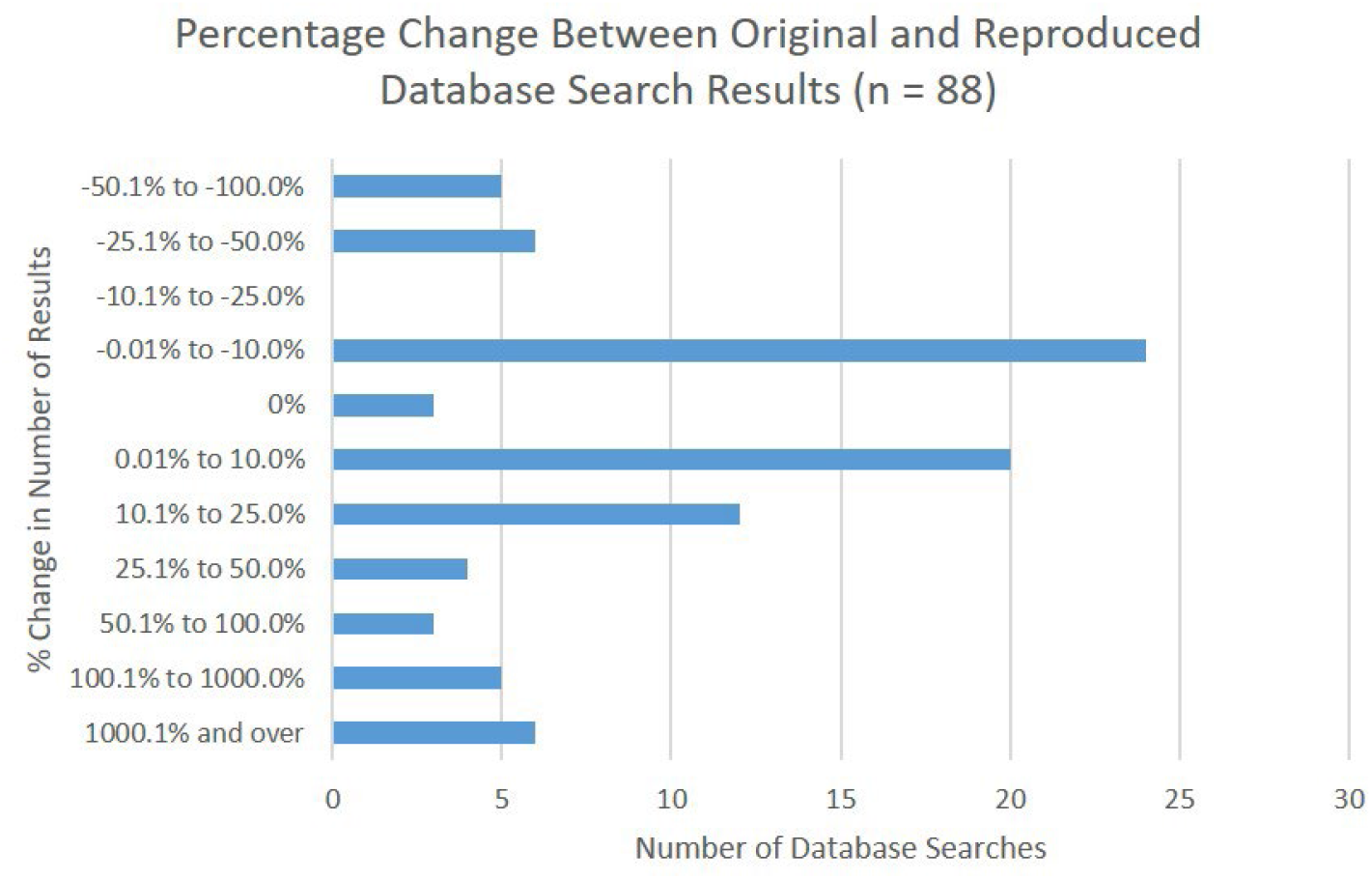
Percentage change between original and reproduced database search results

The post-hoc sensitivity analysis examining the impact of altering the threshold of 10% difference in results showed minor differences in outcomes. A 5% maximum difference reduced the number of reproducible systematic reviews to zero, and a 50% maximum difference increased it to two reproducible systematic review searches (Table S4 (https://osf.io/c3av8)[20]).

### Characteristics of searches: Guidelines and librarian/information specialist involvement

We examined the characteristics of searches to identify commonalities in reproducible and irreproducible searches. Though there were few clear-cut commonalities in either group, some trends were visible. For example, database searches with 10% or less difference fulfilled the criteria for on average 4.9 PRISMA-S items. Database searches where it was impossible to attempt a reproduction only fulfilled the criteria for on average 1.7 PRISMA-S items (Table S5 (https://osf.io/2ztmc)[20]). Librarians or information specialists (LIS) were co-authors on 14.9% (7/47) of searches with 10% or less difference in results, but also co-authored 23.5% (4/17) of database searches that would not run. Similarly, 27.7% (13/47) systematic reviews which we reproduced with 10% or less difference mentioned or acknowledged LIS, but they were also mentioned or acknowledged in 41.2% (7/17) of the seventeen searches which would not run (Figure 3).

**Figure 3.**
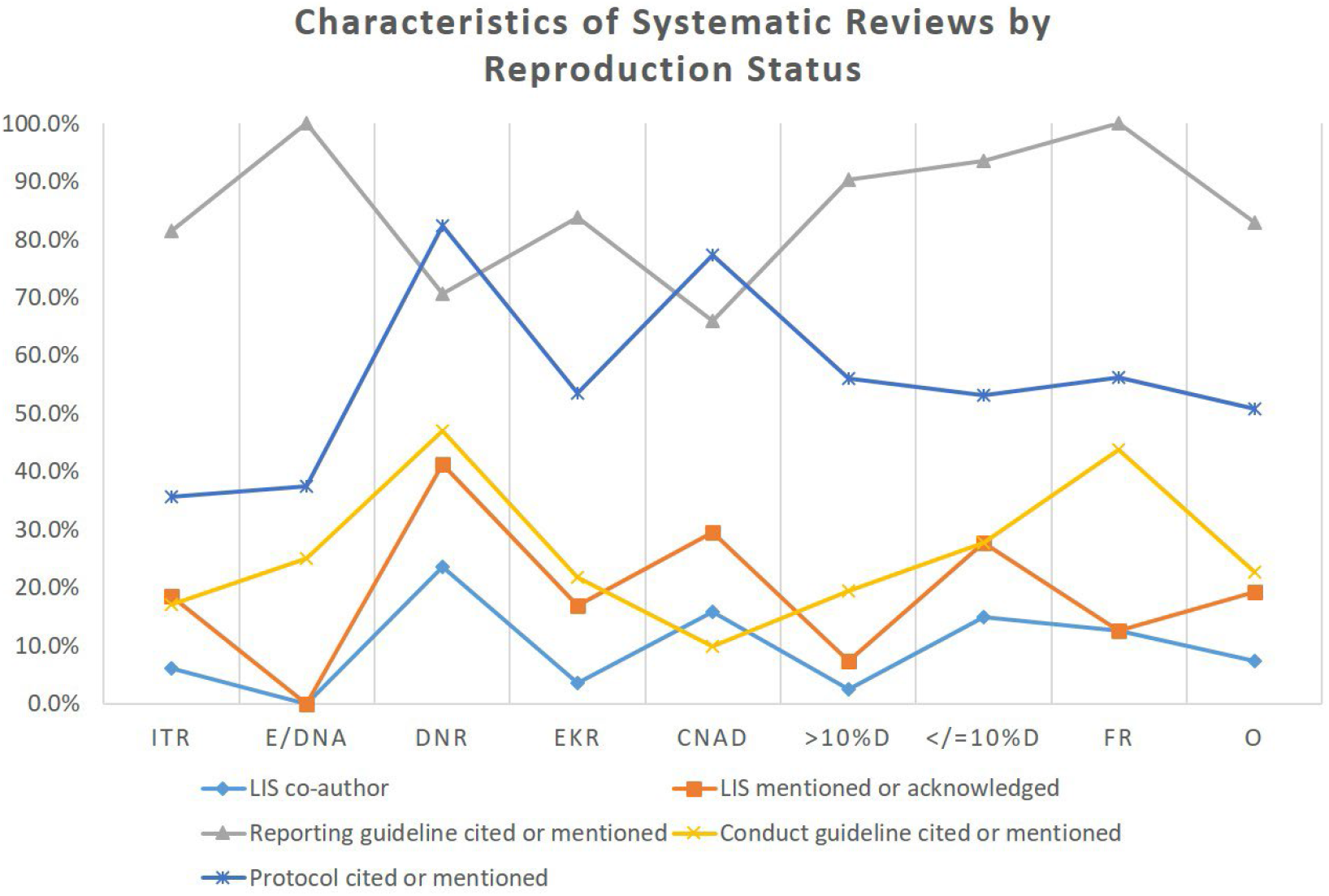
Percentage of database searches with systematic review characteristics by reproduction status. Abbreviations: ITR: Impossible to reproduce (n = 146); E/DNA: Excluded or database not accessible (n = 16); DNR: Did not run (n = 17); EKR: Expert knowledge required (n = 142); CNAD: Could not assess difference (n = 44); >10%D: Greater than 10% difference (n = 41); </=10%D: Less than or equal to 10% difference (n = 47); FR: Fully reproducible (n = 16); O: Overall (n = 453).

### Characteristics of searches: search strategy location and format

The location and format of the search strategies differed between reproducible and irreproducible searches. Fully reproducible database searches and those with a difference less than 10% between the original and the reproduction were located in the supplementary materials the majority of the time (68.8% (11/16) and 71.4% (35/47), respectively). With the exception of two database searches, all other searches in these categories were published in an appendix to the article. For irreproducible searches, there was more variation in where the search or search description was located. None of the systematic reviews in our sample published their search strategies in a repository (Table S5 (https://osf.io/2ztmc)[20]).

Searches that were specific to an individual database were more likely to be reproduced with 10% or less difference in results (Figure 4). Only four (8.5%) of the 47 searches with 10% or less difference were generic, or designed for more than one database search. Comparatively, 48.6% (69/142) of database searches that required expert knowledge to reproduce were generic. 81.3% (13/16) of the fully reproducible searches and 63.8% (30/47) of the searches reproduced with less than 10% difference were multi-line searches, meaning that terms or concepts were searched on separate lines and combined later. Single line searches, on the other hand, were commonly employed in database searches that required expert knowledge to conduct (57.7%; 82/142) and those with greater than 10% difference in results (65.9%; 27/41).

**Figure 4.**
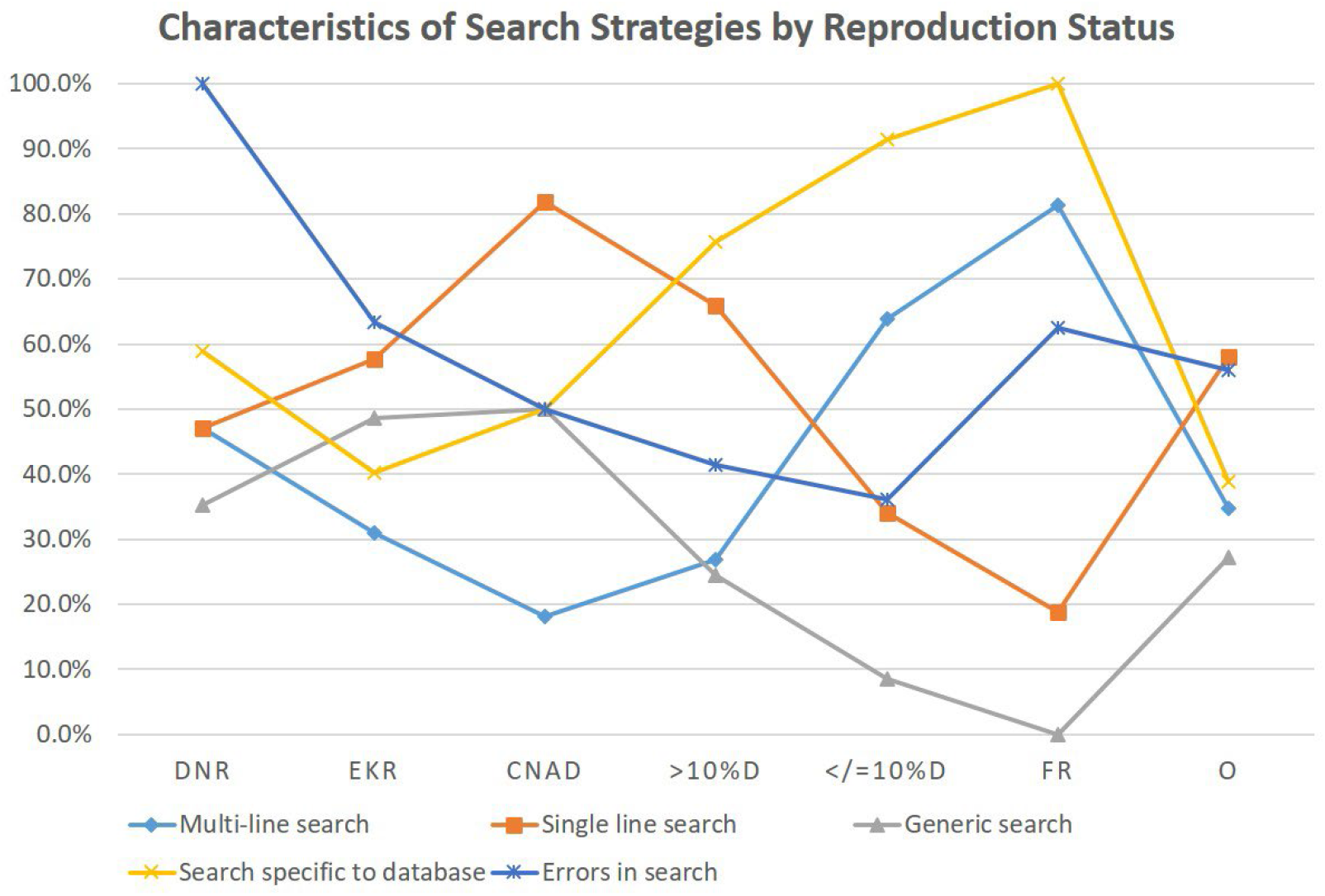
Percentage of database searches with search strategy characteristics by reproduction status. Abbreviations: DNR: Did not run (n = 17); EKR: Expert knowledge required (n = 142); CNAD: Could not assess difference (n = 44); >10%D: Greater than 10% difference (n = 41); </=10%D: Less than or equal to 10% difference (n = 47); FR: Fully reproducible (n = 16); O: Overall (n = 453).

### Characteristics of searches: errors

Overall, 56.0% (163/291) of all database searches contained at least one error. Errors did not necessarily impact the reproducibility of the search. For example, though all 17 database searches that did not run had errors, and indeed major errors that impacted the ability to conduct the search, 62.5% (10/16) of the fully reproducible database searches had errors as well. The lowest prevalence of errors was for database searches with results variance less than 10% (36.2%; 17/47). Unintentional spelling errors were present, but uncommon, occurring in 3.8% (11/291) of the database searches for which reproduction was attempted. Most common were “other” errors, which occurred in 36.1% (105/291) of database searches (Table 2). “Other” errors largely included Boolean logic errors, such as missing parentheses or phrasing, but also included examples of duplicative lines, terms or phrases that were not in the database’s index, mislabeled line numbers, using incorrect or erroneous field codes, and not using all lines, amongst others.

**Table 2.** Types of errors found in published database searches by reproduction status

|  | Did Not Run<br>n = 17 |  | Expert Knowledge<br>Required<br>n = 142 |  | Could Not Assess<br>Variation<br>n = 44 |  | >10% variation<br>n = 41 |  | </=10% variation<br>n = 47 |  | Fully reproducible<br>database searches<br>n = 16 |  | Overall<br>n = 291 |  |
| --- | --- | --- | --- | --- | --- | --- | --- | --- | --- | --- | --- | --- | --- | --- |
|  | n | % | n | % | n | % | n | % | n | % | n | % | n | % |
| **Typographical** | 2 | 11.8% | 26 | 18.3% | 3 | 6.8% | 4 | 9.8% | 8 | 17.0% | 7 | 43.8% | 50 | 17.2% |
| **Spelling** | 1 | 5.9% | 5 | 3.5% | 1 | 2.3% | 0 | 0.0% | 2 | 4.3% | 2 | 12.5% | 11 | 3.8% |
| **Syntax** | 8 | 47.1% | 34 | 23.9% | 16 | 36.4% | 8 | 19.5% | 2 | 4.3% | 2 | 12.5% | 70 | 24.1% |
| **Other** | 13 | 76.5% | 56 | 39.4% | 9 | 20.5% | 9 | 22.0% | 9 | 19.1% | 9 | 56.3% | 105 | 36.1% |

## DISCUSSION

### Principal findings

Only one systematic review search in our sample was fully reproducible, with all database searches for the review meeting six key PRISMA-S reporting criteria and reproduced with 10% or less difference in the number of results between the originally reported search and the reproduction. Our findings show major challenges remain for systematic review search reporting, confirming prior research.[4, 5, 9, 10, 12-15, 29-32] Because our study used PRISMA-S items and attempted to reproduce each search as exactly as possible, our findings are unique and showcase a far grimmer picture of search reproducibility and reporting than has been shown before.[6, 9, 33] Though our findings do not point to a single primary reason for lack of reproducibility, one of the main reasons for irreproducibility is that authors simply do not provide the information required about the databases/platforms they use. Furthermore, peer review and editorial oversight fails to correct the issue.

### Findings in relation to other studies

This study is the first to examine both systematic review and database search reproducibility and to attempt reproduction of all database searches within a systematic review. At present, ours is the only study with a robust estimate of systematic review search reproducibility based not only on proxy measures, but on actual reproduction results. In addition, only two other published studies have looked at PRISMA-S item compliance; neither of those studies used the full criteria of each PRISMA-S item to assess compliance.[34, 35] In addition, both looked at compliance with all sixteen PRISMA-S items, which meant that systematic reviews that didn’t use some types of searching methodology were penalized. Our study, on the other hand, only assessed compliance with the six PRISMA-S items that would be necessary for any systematic review containing at least one database search.

### Study limitations

The small sample size (100 systematic reviews) limits generalizability and limits statistical testing of associations and differences between subgroups (e.g., Cochrane versus non-Cochrane systematic reviews). Reproducing a larger sample of searches would be resource-intensive and, based on prior research, unlikely to produce significantly different results. We were limited by our access to databases and platforms; the reproduction team did not always have the same versions of resources, meaning our reproduction and validation numbers predictably varied.[25, 36] For example, a search conducted in CINAHL, CINAHL with Full Text, and CINAHL Complete, all on the EBSCOhost platform, will produce slightly different results. We included the allowance for 10% difference between original and reproduction results to account for this variability. There is an urgent need for database consistency and stability within and across platforms.

We did not compare whether the same records were retrieved, nor did we analyze whether studies included in the systematic review would differ between the original search and the reproduction. If the included studies would have been impacted, it is still possible that this would not affect the pooled results of the systematic review.[37, 38] Few systematic reviews in our sample contained the full names of the database used, even when the platform name was provided, so our reproductions and validations were based on what we had access to rather than attempting the search in all possible variations. Lastly, we excluded database searches requiring Chinese or Japanese characters, though only one of those database searches could potentially have been reproduced with the data provided.

This study is intended to benchmark systematic review search reproducibility while PRISMA 2020 and PRISMA-S were in their first year since release.[3, 16, 39] Therefore, we acknowledge that the systematic reviews in our samples likely would neither have been subject to the guidance in PRISMA 2020 or PRISMA-S from the journals they submitted to nor would the authors necessarily be aware of updated guidance while their research was in progress. That being said, PRISMA 2020 and PRISMA-S both build on PRISMA 2009’s guidance for search reporting.[3, 16, 39, 40] Interestingly, the one item that achieved the most compliance in systematic reviews was completely new to the reporting guidance, namely tracking the number of results per database (PRISMA-S item 15). When PRISMA 2020 and PRISMA-S become more widely adopted, we would hope that future studies of search reproducibility will find systematic reviews and database searches with more transparent and complete reporting.

### Study implications

As systematic reviews and clinical practice guidelines based upon them continue to proliferate, so does research waste. Since Ioannidis described the “mass production of redundant, misleading, and conflicting systematic reviews,” systematic reviews continue to increase in popularity.[41] Hoffmann et al. documented the growth of systematic reviews, noting that almost 80 systematic reviews were published each day by 2019.[42] From our search to identify systematic reviews indexed in MEDLINE in a single month in 2021, we found approximately 135 systematic reviews per day, a substantial increase in two years. Unfortunately, the improvement in systematic review search transparency or reproducibility seems to trail behind and irreproducible, poorly reported, and poorly conducted systematic reviews continue to be published.[4-7, 33] This directly impacts patients and the public through an influx of irreproducible systematic reviews and clinical practice guidelines.

One of the primary reasons for including full search strategies in systematic review publications is to reduce research waste by enabling the reuse of prior published strategies to update systematic review findings.[1, 43] If searches are not reproducible or require significant expertise to re-execute, there is little point in attempting to update the systematic review. Starting a fresh search from scratch may be the only option in these cases.

Systematic reviews are not the only type of research facing a reckoning with poor reporting and lack of reproducibility.[18, 44] Studies of the reproducibility of shared data and code have similarly shown that, despite journal policies on sharing, published research often remains irreproducible.[45, 46] As calls for data sharing in systematic reviews increase and gain traction,[6, 47] we hope that systematic review searches will be acknowledged as essential data and code to preserve, document, and share.

## CONCLUSIONS

Systematic reviews should be reproducible, but they are not. To correct this will require a multi-faceted response from searchers, systematic review teams and authors, peer reviewers, journal editors, and database providers. Using reporting guidelines as intended, particularly PRISMA-S and PRISMA 2020, can help guide authors and searchers on best practices for transparent reporting.

## Data Availability

All data in this study is publicly available on an OSF Project site (https://dx.doi.org/10.17605/OSF.IO/UGNCT), including raw and summarized data. This includes the results of all data extraction, citations to included and not included studies, and screenshots and documentation on the process of reproducing and validating search strategies. The data is available under a CC-By Attribution 4.0 International license.

## Abbreviations

LIS: Librarian or information specialist
PRISMA: Preferred Reporting Items for Systematic reviews and Meta-Analyses reporting guideline
PRISMA-S: Preferred Reporting Items for Systematic reviews and Meta-Analyses literature search extension

## Author contributorship

MLR conceived the study idea. MLR, SS, LMB, DM, JK, and MPZ designed the study. MLR, JK, and MPZ were responsible for the analysis design and conduct. MLR, TJB, and CP performed data collection and management and finalized data interpretation. MLR, TJB, and CP conducted search reproductions. MLR wrote the first draft of the manuscript. MLR, TJB, CP, SS, LMB, MPZ, JK, and DM contributed to and approved the final manuscript. The corresponding author attests that all listed authors meet authorship criteria and that no others meeting the criteria have been omitted.

## Ethics approval

No ethics approval was required for this study, which is a meta-research study.

## Registration

This study is registered using Open Science Framework (https://doi.org/10.17605/OSF.IO/KBVSR).

## Data availability statement

All data in this study is publicly available on an OSF Project site,[20] including raw and summarized data. This includes the results of all data extraction, citations to included and not included studies, and screenshots and documentation on the process of reproducing and validating search strategies. The data is available under a CC-By Attribution 4.0 International license.

## Acknowledgements

We give special thanks to Patti McCall (Health Science Center Libraries, George A. Smathers Libraries, University of Florida) for her contributions to screening, data extraction, and PRISMA-S adherence evaluation. We also are grateful to John Reynolds (Louis Calder Memorial Library, University of Miami) for his work validating search reproductions. We thank Tom Roper (formerly Brighton and Sussex University Hospitals NHS) for alerting us to the now defunct Healthcare Databases Advanced Search (HDAS) platform that was used in one of the systematic reviews. Finally, we acknowledge the contributions of Nikki Dettmar (University of Washington), Kjell Johnson (Østfold University College), Marte Ødegaard (University of Oslo), and Judy Wright (University of Leeds), each of whom conducted a search reproduction.

## Transparency statement

The lead author affirms that the manuscript is an honest, accurate, and transparent account of the study being reported; that no important aspects of the study have been omitted; and that any discrepancies from the study as originally registered have been explained.

